# Analysis of the mitigation strategies for COVID-19: *from mathematical modelling perspective*

**DOI:** 10.1101/2020.04.15.20066308

**Authors:** S. M. Kassa, H.J.B. Njagarah, Y. A. Terefe

## Abstract

In this article, a mathematical model for the transmission of COVID-19 disease is formulated and analysed. It is shown that the model exhibits a backward bifurcation at ℛ_0_ = 1 when recovered individuals do not develop a permanent immunity for the disease. In the absence of reinfection, it is proved that the model is without backward bifurcation and the disease free equilibrium is globally asymptotically stable for ℛ_0_ < 1. By using available data, the model is validated and parameter values are estimated. The sensitivity of the value of ℛ_0_ to changes in any of the parameter values involved in its formula is analysed. Moreover, various mitigation strategies are investigated using the proposed model and it is observed that the asymptomatic infectious group of individuals may play the major role in the re-emergence of the disease in the future. Therefore, it is recommended that in the absence of vaccination, countries need to develop capacities to detect and isolate at least 30% of the asymptomatic infectious group of individuals while treating in isolation at least 50% of symptomatic patients to control the disease.

## 1 Introduction

A novel coronavirus, named a severe acute respiratory syndrome coronavirus 2 (SARS-CoV-2; previously known as 2019-nCoV), was identified as the infectious agent causing an outbreak of viral pneumonia in Wuhan, China, in December 2019 [43]. The World Health Organization (WHO) medical team codenamed the new outbreak caused by SARS-CoV-2 as “coronavirus disease 2019 (COVID-19)”. The infection is in the same category as the Severe Acute Respiratory Syndrome (SARS) which emerged in Southern China in 2002, spreading to up to 30 Countries, with a total of 8,098 cases and claiming 774 lives [33]. COVID-19 is also in the same category as the Middle East Respiratory Syndrome (MERS) which was first identified in Saudi Arabia in 2012, and ended up spreading to 27 countries around the world, reaching a total of 2,519 cases confirmed and claiming up to 866 lives [41].

Since January 2020, an increasing number of cases confirmed to be infected with COVID-19 were detected outside Wuhan, and currently it has been spread all over the world. As of April 14, 2020, (08:26 GMT), COVID-19 had affected all continents including island nations (210 countries and territories as well as 2 international conveyances), with the total number of cumulative infections globally standing at 1,929,995 cases and 119,789 deaths [48], and the numbers is still increasing.

The major portal of entry of the virus in the body is the tissue lining the T-zones of the face (including the nose, eyes and mouth). The infection is characterised by loss of the sense of smell (a condition referred to as hyposmia/anosmia), taste and poor appetite. Although, such conditions have been observed in COVID-19 patients, many carriers of the infection may not show any severe symptoms like fever and cough but have hyposmia, loss of taste and loss of appetite.

Whereas knowledge of the virus dynamics and host response are essential for formulating strategies for antiviral treatment, vaccination, and epidemiological control of COVID-19, estimation of changes in transmission over time can provide insights into the epidemiological situation and help to identify whether public health control measures are having a measurable effect [5, 35]. The analysis from mathematical models may assist decision makers to estimate the risk and the potential future growth of the disease in the population. Understanding the transmission dynamics of the infection is crucial to design alternative intervention strategies [28]. In general, by approaching infectious diseases from a mathematical perspective, we can identify patterns and common systems in disease function, which would enable us to find some of the underlying structures that govern outbreaks and epidemics.

Mathematical models that analyse the spread of COVID-19 have began to appear in few published papers and online resources [2, 9, 12, 28, 36]. However, there are several challenges to the use of mathematical models in providing nearly accurate predictions at this early stage of the outbreak, particularly in real time as it is difficult to determine many of the pathogen-based parameters through mathematical models. The estimation of such parameters will require clinical observations and “shoe-leather” epidemiology [30]. It is also possible for some of the parameters to be verified through observation only at a later stage of the course of the epidemics. To get better predictions and to design and analyse various alternative intervention strategies in the absence of such parameter values, one needs to estimate them from existing epidemiological data.

Like many respiratory viruses, the novel coronavirus SARS-CoV-2 can be spread in tiny droplets released from the nose and mouth of an infected person. As soon as the virus enters the body (either through the mouth, nose or membrane of the eyes), it finds its way to the windpipe and then the lungs. This viral attack is characterised by flu like symptoms, fever (body temperatures > 38.3°C) and dry cough at the initial stage of the infection. Once the virus gets into the lungs, it causes fibrosis of the lungs leading to shortness of breath (or difficulty to breath) and severe pneumonia followed by impaired functioning of the liver and acute kidney injury [20]. The virus is then released from the infected individual when they cough, sneeze and when they touch their own nose or mouth. Some part of the particles that are released through coughing or sneezing may land on clothing of other people in close proximity, and surfaces around them while some of the smaller particles can remain in the air for some times. In addition, some scientific evidences show that the virus can also be shed for a longer time in faecal matters [45]. The virus survives on surfaces, fabrics, metals, plastics for variable times. Recent report indicates that the virus can survive from shorter time (in the air) up to 2-3 days long on plastic and stainless-steel surfaces [39]. This implies that an uninfected individual can also acquire the virus through contact of infected surfaces.

That means, there is a possibility for COVID-19 infection to spread from such contaminated surfaces and objects to uninfected humans. Hence, including the proportion of indirect transmission from the environment in the mathematical model structure is important to address this situation. We note that the impact of environmental contamination and its role in the transmission of the disease is not well studied in mathematical models developed so far.

Reinfection by the family of coronavirus is possible as it is indicated in [21, 50]. Even if it is not yet well known how long it takes for a person who recovered from COVID-19 to loose immunity, we can not overlook its impact at this stage. Therefore, when formulating a mathematical model for COVID-19 dynamics, it is reasonable to consider a kind of Susceptible-Infected-Recovered-Susceptible (SIRS) type of epidemiological model formulation.

So far there is no known curing medicine nor vaccine to combat the COVID-19 pandemic. The available prevention mechanisms that are recommended by the WHO so far are also limited and their effectiveness is not yet fully tested. The population level application of these preventive mechanisms varies from region to region and from country to country. In some places, the protective measures are employed voluntarily by individuals and in some other places governments impose some kind of rules on the population to use strict social distancing and wearing face masks at public places. However, the adherence to these rules is not uniform.

In the past, it has been witnessed that during the outbreak of infectious diseases, the human population has been taking precautionary actions such that wearing masks, abstinence from risky contacts, avoiding public transport means and increasing the uptake of vaccination (when available) [14, 24, 32]. Behaviour change towards using preventive mechanisms by the population to protect themselves from an infectious disease is assumed to be dependent on the way that the disease is transmitted and its fatality. Individuals who have awareness about the disease and who decided to use preventive mechanisms have less susceptibility than those without awareness and demonstrating the usual risky behaviour.

A number of mathematical models have been proposed to analyse the effects of human behaviour in the dynamics of infectious diseases (see [13, 15, 17, 22, 25, 26, 29, 32, 40], and the references therein). In this paper, we follow the diffusion of innovation approach, which was proposed by Kassa and Ouiniho [24]. In models of this approach, it is assumed that the perceived threat for the population is the level of prevalence of the disease. However, for diseases with short time cycles, the prevalence dependent awareness function looks non-realistic. Therefore, in this work we assumed that awareness is driven by the magnitude of the incidence rate reported each day. That means, based on the diffusion of innovation method one may consider the *perceived threat* for the population to be the incidence of the disease.

At the beginning of the COVID-19 outbreak, a huge disparity has been observed in the use of self-protective mechanisms and adherence to the advises given by public health agencies. In particular, the people in some parts of Asia have fully embraced the measures while many in other parts of the world were very much hesitant to use them. For example, wearing a face mask every day in public appearance is like a ritual in most of the countries in South East Asia, while the same is considered as a bad gesture in many of the other parts of the world [46] (even if it is now becoming a “normal norm” also everywhere in the globe). One key difference between these societies and the people in the West, is that the communities in the South East Asia have experienced similar disease outbreaks before and the memories are still fresh and painful [46]. That means, recent history of a similar event plays a role in behavioural change of the population specially at the beginning of the outbreak in addition to the perceived threat from the disease.

Therefore, in this paper, we consider a mathematical model that takes into account

1. the transmission dynamics of COVID-19 similar to the SIRS model,
2. the contribution of the asymptomatic infectious individuals in the transmission dynamics of the disease in the population,
3. the effect of indirect transmission of the disease through the environment,
4. behavioural change of individuals in the society to apply self-protective measures, and
5. the intensity of historical events from recent similar outbreaks.

By analysing the proposed mathematical model, the effect of each of these factors is investigated in terms of their contribution in the control strategies of the disease.

The layout of this paper is as follows: The model is described and formulated in the next section. Its qualitative analysis is presented in Section 3. Estimation of the parameters and the sensitivity analysis of the reproduction number of the model with respect to involved parameters are discussed in Section 4. Numerical simulations of the model with some assumed intervention scenarios are also presented in this same section. Concluding remarks of the study are given in Sections 5.

## 2 Model formulation

In this section, we present a mathematical model for the transmission dynamics of COVID-19 which spreads in a population. The susceptible individuals can be infected through either direct contact with infectious individuals or indirect contact with novel coronavirus infected environment. The population under consideration is grouped into disjoint compartments. Individuals who are susceptible to the disease and without formal awareness about the prevention mechanisms or who did not decide to use any one of them are grouped in the *S* class. Individuals who are susceptible but are aware of about and decided to apply any of the existing protective mechanisms after receiving public health information on how to protect themselves from the novel coronavirus infection are placed in the *S*_*e*_ class. COVID-19 infected individuals who are asymptomatic and symptomatic are grouped in classes *C* and *I*, respectively. Some studies consider the asymptomatic class as the “exposed” class (see for instance [9]). But since the individuals in this group are known to be infectious and some of them also recover from the disease without going through the *I* group [8], we used a name “carrier” to avoid confusion. The *R* class contains the recovered individuals from COVID-19. Finally, *E* denotes the amount of the novel coronavirus pathogen that contaminates the environment due to shedding by COVID-19 infectious individuals. In the analysis of the model, we intentionally excluded the actual exposed class for mathematical simplicity. However, a 5 days incubation period is taken into consideration in the numerical simulation part of this paper.

By combining the direct and indirect way of transmissions, the force of infection is assumed to have the form

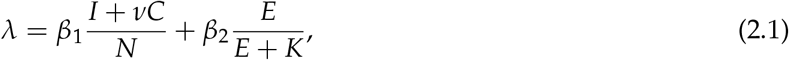

where *N* = *S* + *S*_*e*_ + *C* + *I* + *R* and *K* is the concentration of the novel coronavirus in the environment which increases 50% chance of triggering the disease transmission.

The proposed flow diagram for the transmission dynamics of COVID-19 is depicted in Figure 1 while the description of each of the state variables is given in Table 1.

**Table 1:** Description of the model variables

| Variables | Description |
| --- | --- |
| $S$ | Susceptible population. |
| $S_e$ | Susceptible individuals who are educated to prevent the disease. |
| $C$ | Carriers individuals (infected & infectious but asymptomatic). |
| $I$ | Infected individuals (symptomatic). |
| $R$ | Recovered individuals. |
| $E$ | Contaminated surfaces or objects in the environment. |

**Figure 1:**
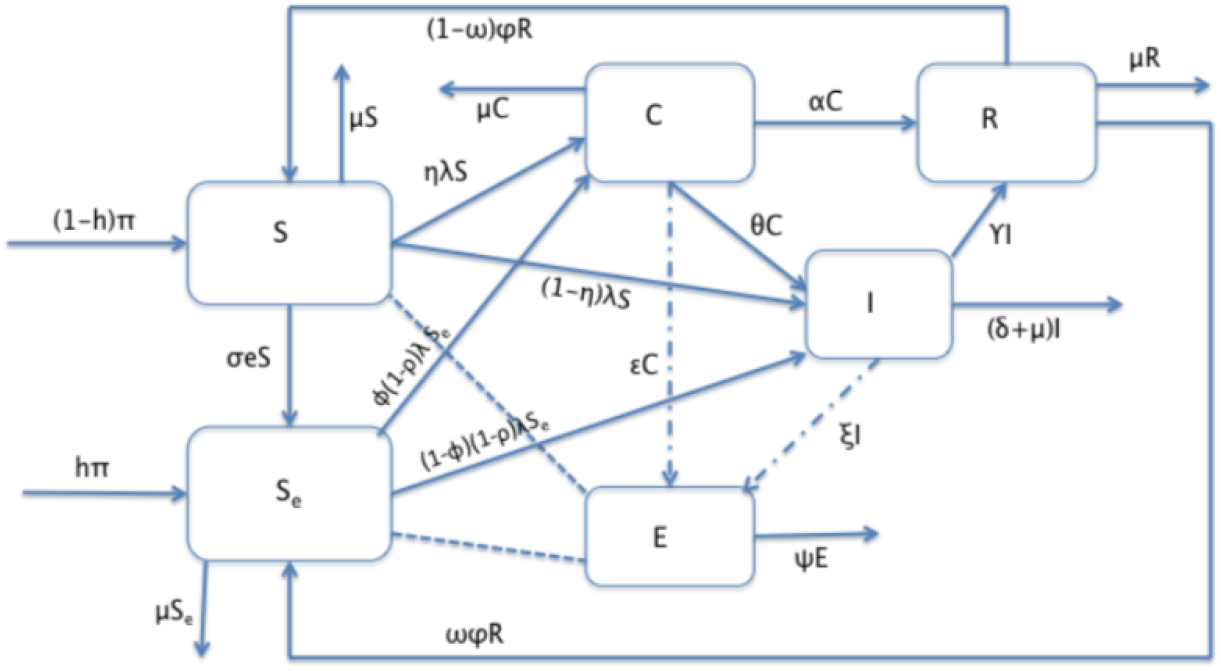
Schematic diagram of the proposed model.

The dynamics of the pandemic is described by using the following system of differential equations (see Table 2 for the description of the involved parameters):

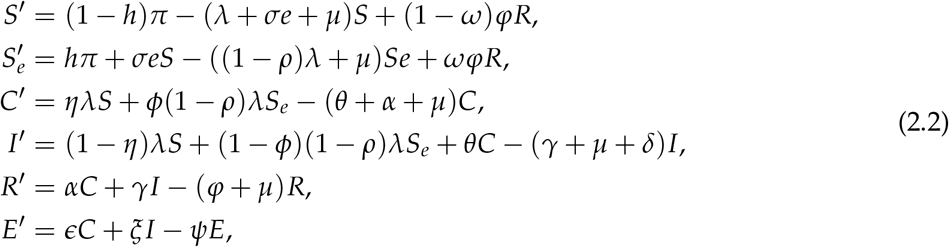

where

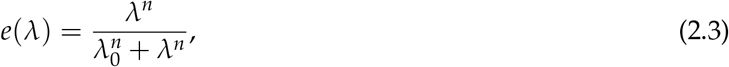

with *λ*_0_ is the value of the force of infection corresponding to the threshold infectivity in which individuals start reacting swiftly (that means, the point at which the behaviour change function changes its concavity). We appended the following nonnegative initial conditions with the system (2.2):

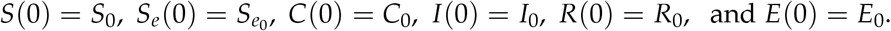

**Table 2:**
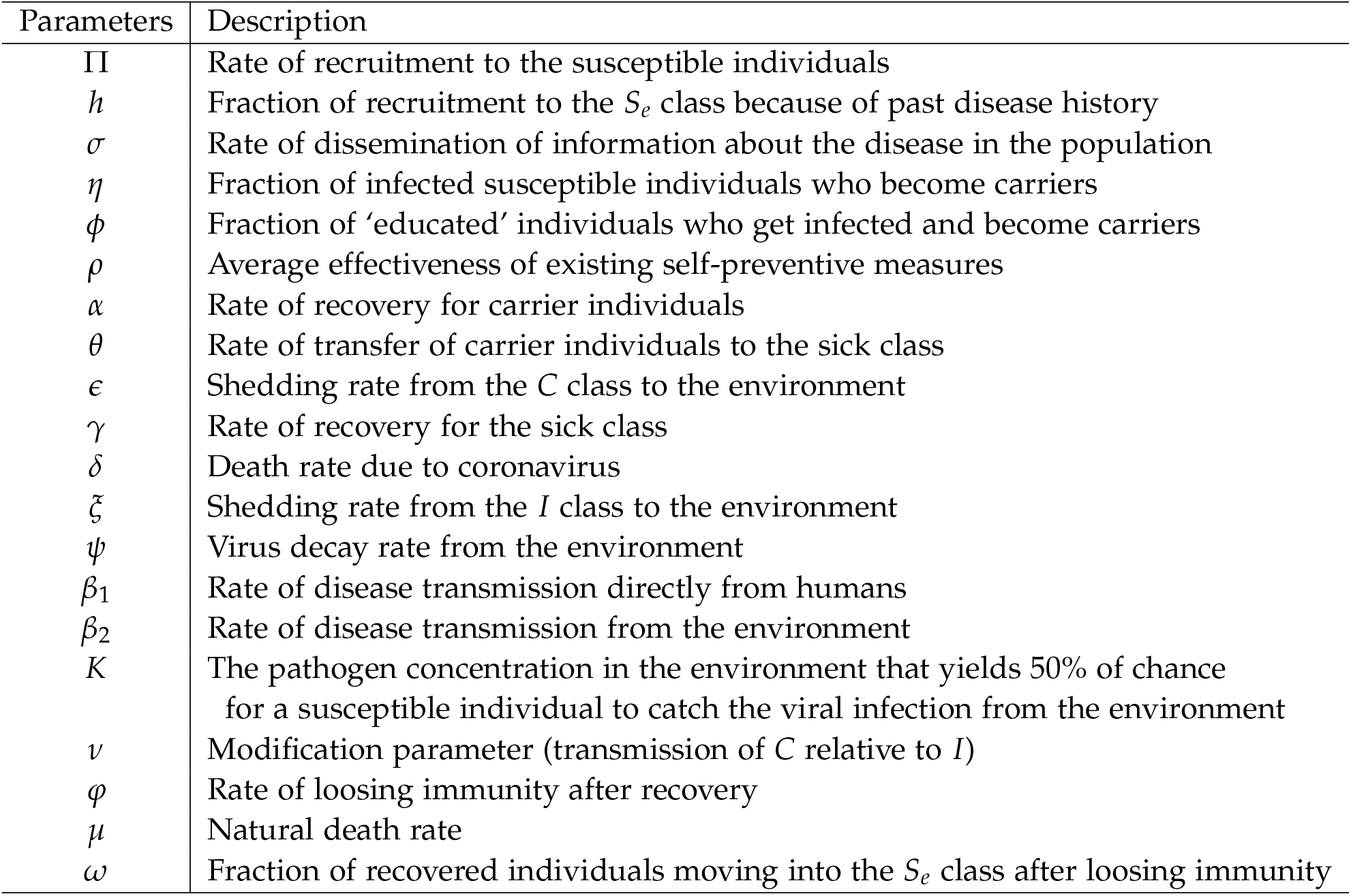
Description of the model parameters

## 3 Analysis of the model

In this section, we study the quantitative and qualitative analysis of the model system Eq. (2.2).

### 3.1 Well-posedness

We begin by determining the biologically feasible set for the model (2.2). The following theorem implies that the solutions of (2.2) are nonnegative and bounded from above, provided that the initial conditions are nonnegative.

#### Theorem 3.1

*Equation (2*.*2) defines a dynamical system on* Ω, *where*

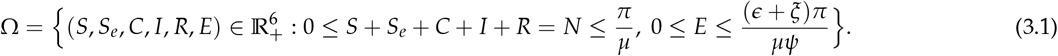

**Proof:** The proof of this Theorem is outlined in Appendix A

### 3.2 Asymptomaticstability of the disease-free equilibrium

To determine the equilibrium solutions, we set the right-hand side of (2.2) equal to zero and obtain

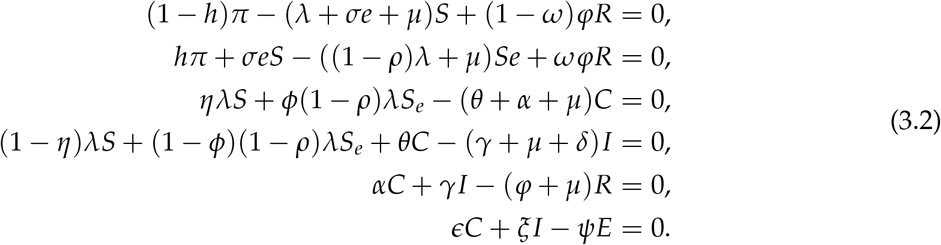

Then, the disease-free equilibrium (DFE) is obtained to be

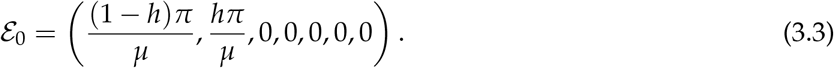

The basic reproduction number, which is very important for the qualitative analysis of the model, is determined here below by using the method of the next generation matrix used in [11, 38]. For the model under consideration, using the notation *X* = (*C, I, E*), we have the vector functions

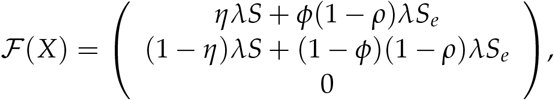

and

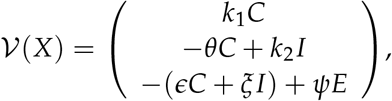

with *k*_1_ = *θ* + *α* + *µ* and *k*_2_ = *γ* + *µ* + *δ* represent the rates at which the disease compartments increase and decrease in size due to the infection, respectively. Then the next generation matrix is

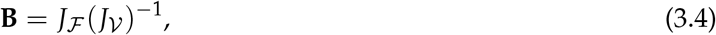

where

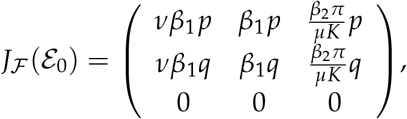

and

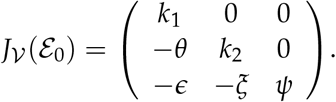

where *p* = *η*(1−*h*) + *ϕ*(1−*ρ*)*h*, and *q* = (1−*η*)(1−*h*) + (1−*ϕ*)(1−*ρ*)*h*. Here, it is not difficult to show that

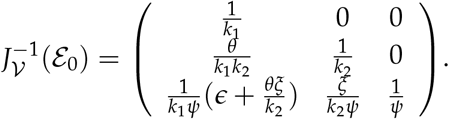

The basic reproduction number denoted by ℛ_0_ is defined as the average number of secondary cases produced in a completely susceptible population by a typical infected individual during its entire period of being infectious [11, 38]. Mathematically, ℛ_0_ is the spectral radius of **B** in Eq. (3.4) and after further simplification, we obtain

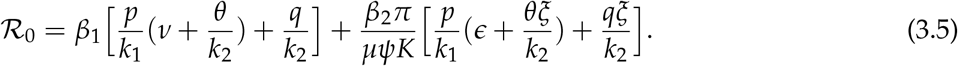

The next result is a direct application of Theorem 2 in [38].

#### Theorem 3.2

*The DFE* E_0_ *of the model (2*.*2) is locally asymptotically stable whenever* ℛ_0_ < 1 *and unstable if* ℛ_0_ > 1.

The epidemiological implication of Theorem 3.2 is that the transmission of COVID-19 can be controlled by having ℛ_0_ < 1 if the initial total numbers in each of the subpopulation involved in Eq.(2.2) are in the basin of attraction of *ε*_0_. To ensure that eliminating the disease is independent of the initial size of the subpopulation, the disease-free equilibrium must be globally asymptotically stable when ℛ_0_ < 1. This is what we present here below.

#### Theorem 3.3

*The model (2*.*2) undergoes a backward bifurcation at* ℛ_0_ = 1 *when the parameters satisfy the condition*

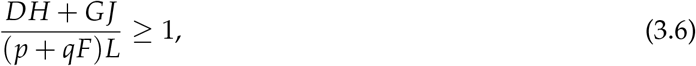

*where*

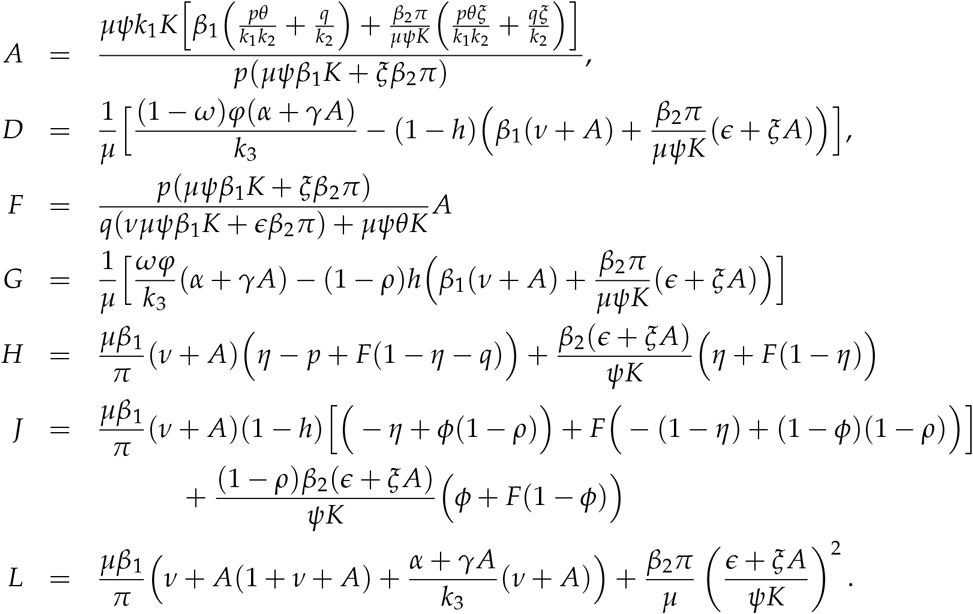

The proof of Theorem 3.3 is carried out using the center manifold theory in [6] and is given in Appendix B

In the above theorem (Theorem 3.3), if the reinfection parameter *ϕ* = 0, we can observe that both *D* and *G* are negative. Hence, Eq. (3.6) can not be true. In this case, we give below a Theorem which asserts the global stability of the DFE of the model.

#### Theorem 3.4

*The disease-free equilibrium of system (2*.*2) in the case when ϕ* = 0 *is globally asymptotically stable for* ℛ_0_ < 1.

Proof: To prove the theorem for the case *ϕ* = 0, we use Kamgang-Sallet Stability Theorem stated in [23]. Let *X* = (*X*_1_, *X*_2_) with *X*_1_ = (*S, S*_*e*_, *R*) ∈ ℝ^3^ and *X*_2_ = (*C, I, E*) ∈ ℝ^3^. Then the system (2.2) can be written as

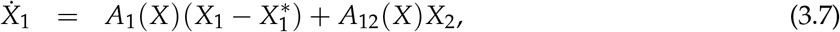

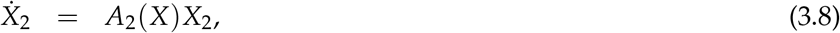

where 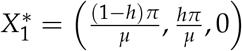,

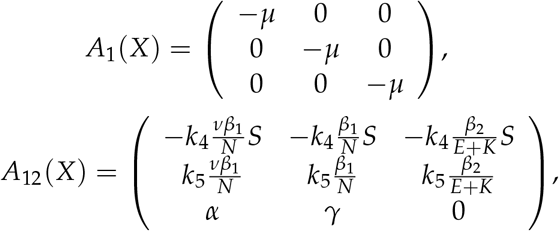

and

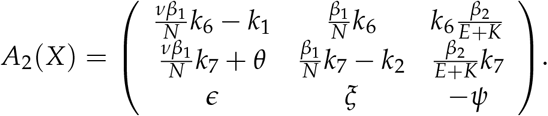

with 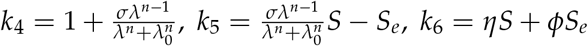 and *k*_*7*_= (1 − *η*)*S* + (1 − *ϕ*)*S*_*e*_.

We show that the five sufficient conditions of Kamgang-Sallet Theorem (in [23]) are satisfied as follows.

1. The system (2.2) is a dynamical system on Ω. This is proved in Theorem 3.1.
2. The equilibrium 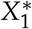 is GAS for the subsystem 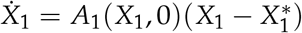. This is obvious from the structure of the involved matrix.
3. The matrix *A*_2_(*X*) is Metzler (i.e., all the off-diagonal elements are nonnegative) and irreducible for any given *X* ∈ Ω.
4. There exists an upper-bound matrix *Ā*_2_ for the set

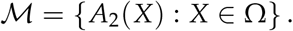

Indeed,

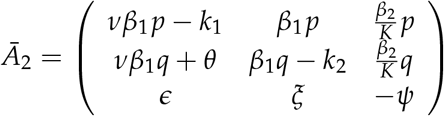

with *p* = (1 − *h*)*η* + *hϕ* and *q* = (1 − *h*)(1 − *η*) + *h*(1 − *ϕ*) is an upper-bound for ℳ. 5. For ℛ_0_ ≤ 1 in Eq. (3.5)

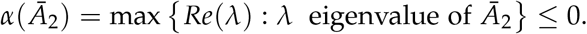

Hence, by the Kamgang-Sallet Stability Theorem [23], the disease-free equilibrium is globally asymptotically stable for ℛ_0_ < 1. □

The reality behind Theorem 3.4 is that, if immunity is permanent (*ϕ* = 0), coronavirus will be effectively controlled in the community if ℛ_0_ can be brought to a value less than unity.

## 4 Numerical Simulations and Discussion

### 4.1 Estimation of parameters from data and literature

The novel coronavirus being a new strain of corona viruses, information about the dynamics of the infection is still evolving. Biological studies of parameter values describing the vital dynamics of the infection are still ongoing as more laboratory test become available. Although some studies have been done on the early dynamics of the disease most especially on data from Wuhan, extensive reading reveals that some of the disease dynamics parameters are highly variable and some process are not fully explored. In this work, we use new cases data from Hubei Province of China extracted from WHO situation reports 1-57 [43], i.e. for the period January 21, 2020 to March 17, 2020. We ought to fit the proposed model to the extracted data and estimate the unknown parameters.

The total population of Hubei province was estimated as 59.2 million. The life expectancy of Hubei province varies depending on the area of dwelling (i.e urban or rural) as well as gender [44]. For urban dwellers, the average life expectancy is estimated to be 75.68 years (with an average being 73.72 years for men and 77.79 years for women). The life expectancy of China of the year 2019 was estimated to be 76.79 years where as that for the year 2020 is estimated as 76.96 years. [31]. Owing to the negligible difference in the provincial and Country wide value, we use the Country life expectancy for the year 2019 which gives an average mortality rate of *µ* = 3.57 × 10−5 per day. The recruitment rate is thus given as *π* = *µ* × *N*_0_, where *N*_0_ is taken to be the total population size, 59.2 million.

The average time period taken for symptoms to appear after exposure is observed to vary considerably with a range between 2-14 days [7], 2-24 days [47], with some outliers going to up to 27 days. The observed median incubation period was nearly 5 days [16]

The time to recovery from the onset of symptoms varies depending on the seriousness of the infection with individuals presenting mild illness observed to recover in an average period of 2 weeks while those presenting serious/critical illness recovering in about 3 to 6 weeks. For our parameter estimate simulations we consider a nominal value of 0.0476 day^−1^ (corresponding to 3 weeks) estimated from an interval (0.0238, 0.0714). We note that a patient is considered recovered: (1) if two swab tests taken in a time interval of at least 24 hours both test negative, (2) if the time taken for after the end of the respiratory symptoms and fever is at least 72 hours.

The waning of the immunity after recovery is estimated to range between 4 months to 1 year, which gives an interval for *ϕ* as (0.00274, 0.00824) day^−1^. For our simulation, we consider a nominal value of *ϕ* = 0.00274 day^−1^ (approximately one year). We propose that at the end of the epidemic, at least 20−65% of the recovering population will learn from the experiences during the infection and even when the acquired immunity wanes, such individuals will become susceptible individuals with past history/knowledge of the disease.

The rate of recovery for the symptomatic individuals (*γ*) in Wuhan varied considerably but majority of individuals who recovered from the virus were discharged from hospital after 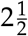 weeks [4]. However, the patients in Wenzhou-China stayed in hispital for 27 days (0.037 per day) on average. In the model fitted on the early trends data from Wuhan-China [27], the recovery rate obtained for symptomatic cases was 0.0897 per day (accounting for 10 days to recovery). The rate of recovery (*α*) for carrier individuals is expected to be higher [9].

According to the WHO situation report [42], it is estimated that up to 80% of COVID-19 cases are asymptomatic or show mild symptoms, 15% show severe symptoms and up-to 5% end up with critical infection and require oxygen or a ventilator. The proportion of individuals who do not show symptoms or have mild symptoms can be as high as 94% [18]. For our model fitting, we use a range of (0.65, 0.86) for both *η* and *ϕ* with selected initial values within the prescribed interval.

Although Hubei province was put on a lockdown on January 23, 2020, the first major decline in the number of new confirmed cases was only observed on February 20, 2020 (Situation report 31 [43]), approximately 1 month after the lockdown was imposed. From February 14, 2020, the method of identification of new cases was revised to include both cases confirmed through laboratory test and clinical observation. As such there was an observed spike in the number of new cases on February 14, 2020 of 4823 compared to 1508 cases (reported on February 13, 2020) and 2420 cases (reported February 15, 2020).

Applying the above described set of assumptions in the bound for some of the parameters, we optimize the model output to fit the daily new cases data reported for the Hubei province, China. The parameter values that best fit to the incidence data is given in Table 3. Figure 2 shows the plot of the reported new-case data together with the incidence of the disease obtained from the model. As we can observe from the graph, the model slightly overestimates the reported data except for the two highest points. In addition, since our model does not assume any control measure at this stage while the reported data after the 31st day may represent the effect of the strict lockdown measure taken by the authorities, the parameters estimated seem to give a good result. When we calculate the value of ℛ_0_ from Eq. (3.5) using the estimated parameters given in Table 3, we obtain ℛ_0_ ≈ 2.91, which is within the range of values reported in [10, 49].

**Table 3:**
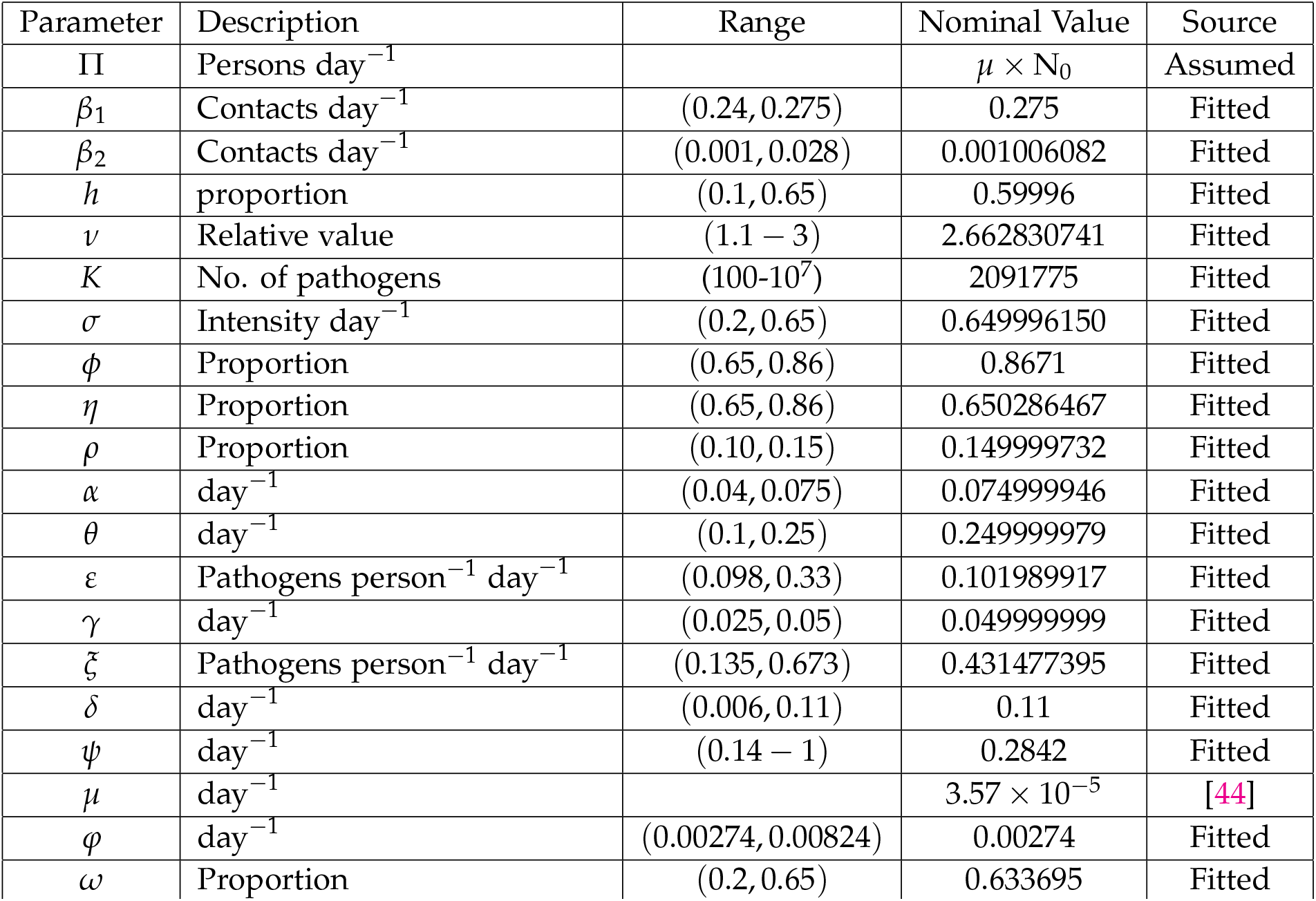
Nominal values and ranges of parameters values.

**Figure 2:**
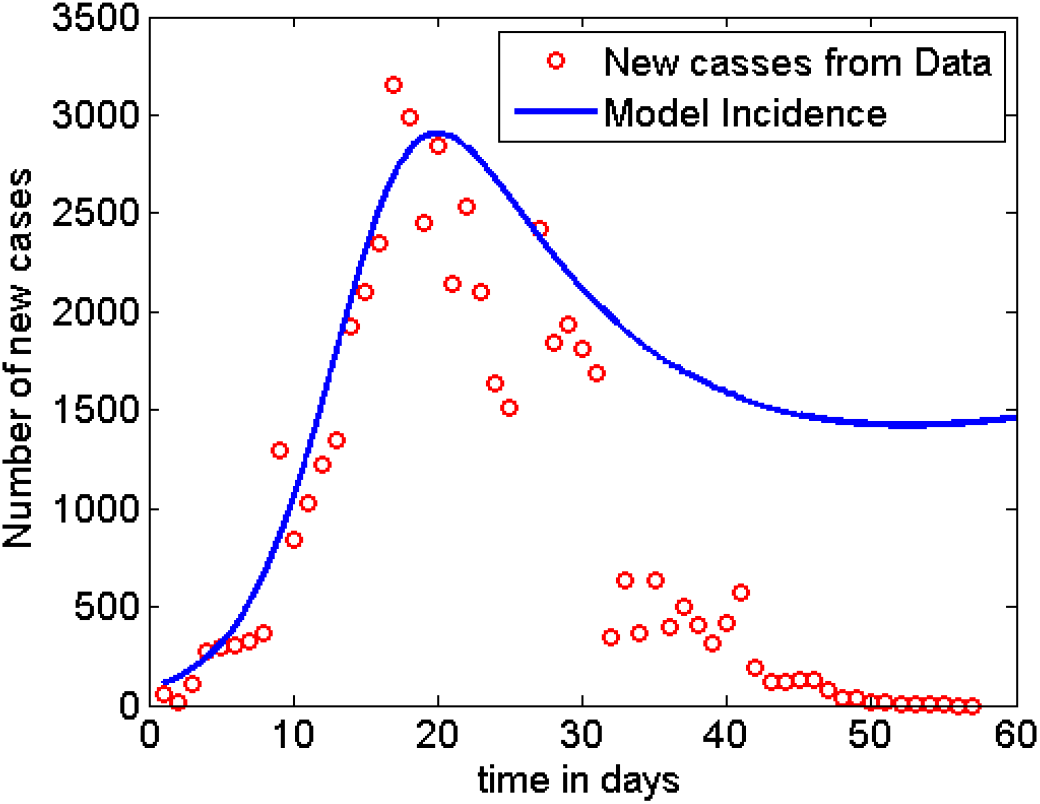
Model fit to the data.

### 4.2 Sensitivity analysis

We examine the sensitivity of ℛ_0_ to variations in parameter values and establish the significance of the sensitivity indices. We used the Latin hypercube Sampling (LHS) scheme, which is an efficient stratified Monte Carlo sampling that allows for simultaneous sampling of the multi-dimensional parameter space [19]. For each run, 1000 simulations were done and Partial Rank Correlation Coefficients (PRCCs) [1] calculated between each of the selected input parameters and the disease threshold. The PRCCs indicate the degree of effect each parameter has on the outcome, which in this case is the disease threshold. The sign of the PRCC identifies the specific qualitative relationship between the input parameter and the output variable. The positive value of the PRCC of the variables implies that when the value of the input parameter increases, the future number of cases will also increases. On the other hand, processes underlying the parameters with negative PRCCs have a potential to contain of the number of cases when enhanced. The results of sensitivity analysis are indicated in Figure 3(a) and the box plot (Figure 3(b)) gives the five-number summary for the computed disease threshold value from the sampled parameter space.

**Figure 3:**
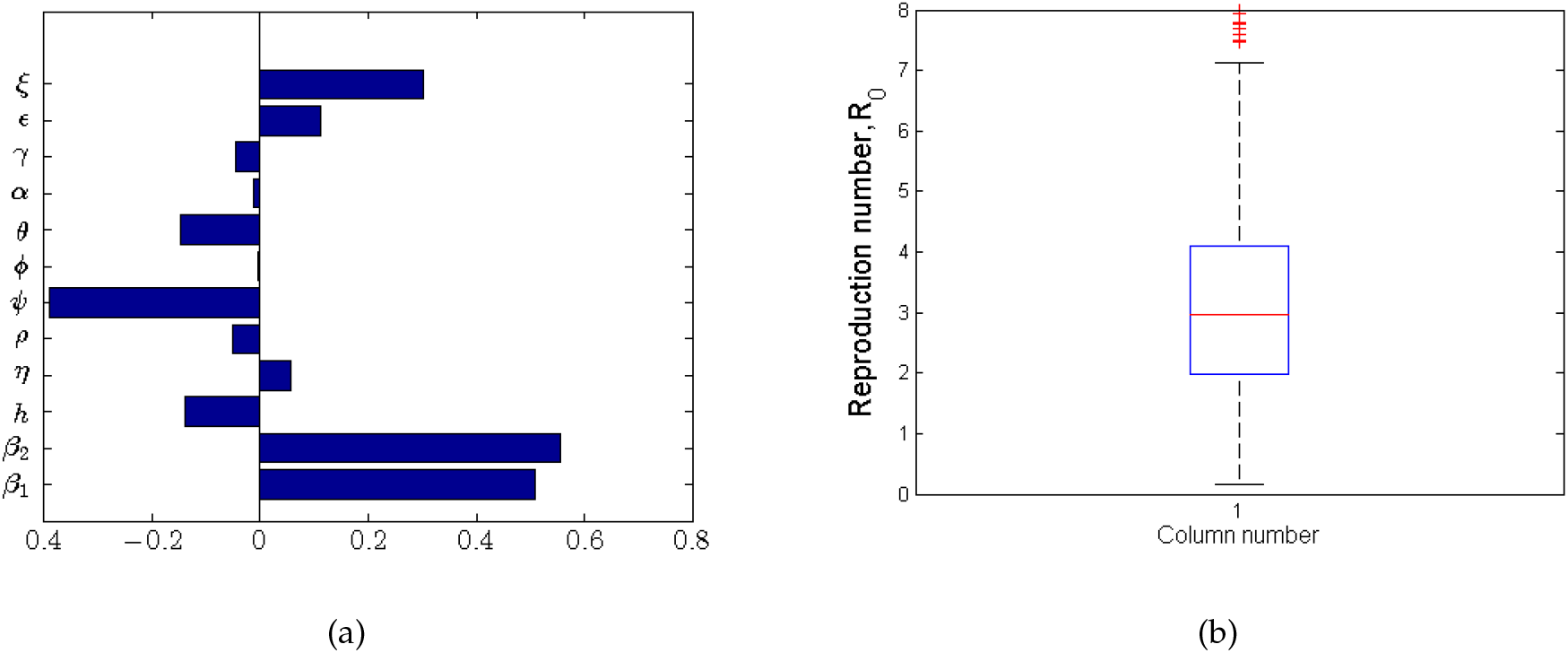
Partial Rank Correlation Coefficients (PRCCs) for a selected range of model parameters in Table 3. The processes underlying the parameters *β*_1_, *β*_2_, *ε* and *ξ* have the greatest potential of making the epidemic worse if increased, whereas processes described by *ψ* and *h* have the greatest potential of containing the epidemic when enhanced.

The processes described by parameters *β*_1_, *β*_2_, *ε* and *ξ* with the greatest positive PRCCs have the greatest potential of worsening pandemic when increased. On the other hand, parameters (*h* and *ψ*) with negative PRCCs have the greatest potential in helping contain the infection when maximised. In this respect, we note that increasing social/physical distancing directly reduces *β*_1_ as this lowers the likelihood of a susceptible individual getting in contact with a potentially infected individual. In addition, practising good hygiene (such as regularly washing hands, using sanitisers to disinfect the infected environment and avoiding touching the T-zones of the face) is associated with lowering the likelihood of contracting the virus from infected surfaces. Anything contrary to the above increases the likelihood of getting the infection through the two aforementioned routes. We further note that practising good hygiene also involves the infected individuals reducing the shedding of the virus into the environment. It is evident from the results in figure 3 and Table 4 that reducing the rate at which the virus is shed into the environment is significant in reducing the severity of the problem.

**Table 4:**
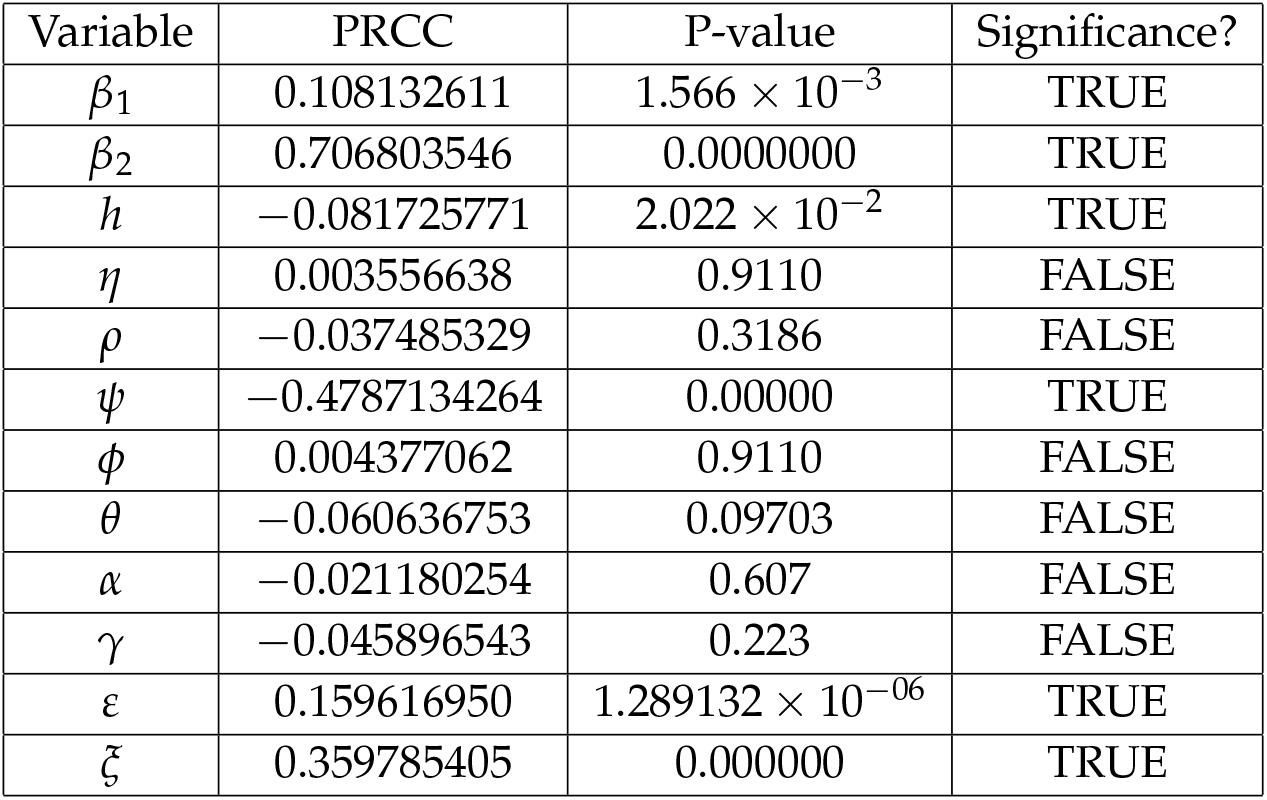
Parameter PRCC significance (FDR Adjusted P-values)

From the five number summary of the results in Figure 3(b), the lower quartile of the computed values of ℛ_0_ is about 2, the median around 2.9 and the upper quartile of about 4. The obtained value of ℛ_0_ is within the range of 3.11(95%*CI*, 2.39 − 4.13) obtained in the early studies in [34]. We note that for a selected combination of underlying processes much higher values of ℛ_0_ can be obtained, which is an indication of possible worsening of the situation. In a similar way, we observe that for particular underlying processes (a selected combination of parameters) the value of ℛ_0_ can be reduced to values below one.

As indicated in [1], we note that although some parameters in the model may have very small magnitudes of PRCCs (non-monotonically related to the disease threshold output), they may still produce sizeable changes in the disease burden. To identify the most important parameters in containing or aggravating the epidemic, we computed p-values for the simulated parameters using Fisher’s Transformation [1]. We note that the computed PRCCs are bounded between the interval [−1, 1]. For this matter, some sampling distribution of variables that are highly correlated is skewed. The Fisher’s Transformation 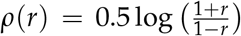 is used to transform the skew distribution to a normal distribution and then compute p-values for each of the parameters based on the PRCCs [1]. The PRCCs for the parameters together with their corresponding p-values are indicated in Table 4.

We carry out pairwise comparison of the significant parameters (whose p-values are less than 0.05, see Table 4) to ascertain whether the process described by such parameters are different. We computed the p-values for the different pairs of significant parameters while accounting for the false discovery rate (FDR) adjustment and the results are given in Table 5.

**Table 5:** Pairwise PRCC Comparisons (FDR Adjusted P-values)

| | $\beta_1$ | $\beta_2$ | $h$ | $\psi$ | $\varepsilon$ | $\xi$ |
| --- | --- | --- | --- | --- | --- | --- |
| $\beta_1$ | | 0 | $2.347 \times 10^{-5}$ | 0 | 0.2443 | $2.642 \times 10^{-9}$ |
| $\beta_2$ | | | 0 | 0 | 0 | 0 |
| $h$ | | | | 0 | $8.647 \times 10^{-8}$ | 0 |
| $\psi$ | | | | | 0 | 0 |
| $\varepsilon$ | | | | | | $1.942 \times 10^{-6}$ |
| $\xi$ | | | | | | |

The major question posed at this point is: Are the different pairs of significant parameters different after FDR adjustment? Based on the FDR adjusted p-values in Table 5, the compared pair of parameters are rendered to be different if their p-value is less than 0.05 and not different otherwise. We summarise our results in Table 6, where “TRUE” indicates that the compared parameters are significantly different and “FALSE” indicating that the parameters are not significantly different.

**Table 6:** Are the parameters different after FDR adjustment?

| | $\beta_1$ | $\beta_2$ | $h$ | $\psi$ | $\varepsilon$ | $\xi$ |
| --- | --- | --- | --- | --- | --- | --- |
| $\beta_1$ | | TRUE | TRUE | TRUE | FALSE | TRUE |
| $\beta_2$ | | | TRUE | TRUE | TRUE | TRUE |
| $h$ | | | | TRUE | TRUE | TRUE |
| $\psi$ | | | | | TRUE | TRUE |
| $\varepsilon$ | | | | | | TRUE |
| $\xi$ | | | | | | |

We observe that the more sensitive parameters are also significantly different (see Table 6) except for the *β*_1_ − *ε* pair which may not necessarily be related.

We examine effect of variation of the sensitive parameters on the reproduction number (ℛ_0_). The results of the variation of parameters with more negative PRCCs are indicated in the bar graphs in Figure (4).

From Figure 4, it is evident that the decay of the virus from the environment (Figure 4(b)) which can be accelerated by disinfecting surfaces reduced the value of ℛ_0_ and consequently the disease burden. In addition, we observe that an increased proportion of individuals with knowledge of similar infections from the past that are practising self-protection and preventive measures (see Figure 4(a)) is important in slowing down the infection at the initial stage. Such proportions of individuals would normally have knowledge about prevention and control mechanisms of the infection just at the onset of the disease.

**Figure 4:**
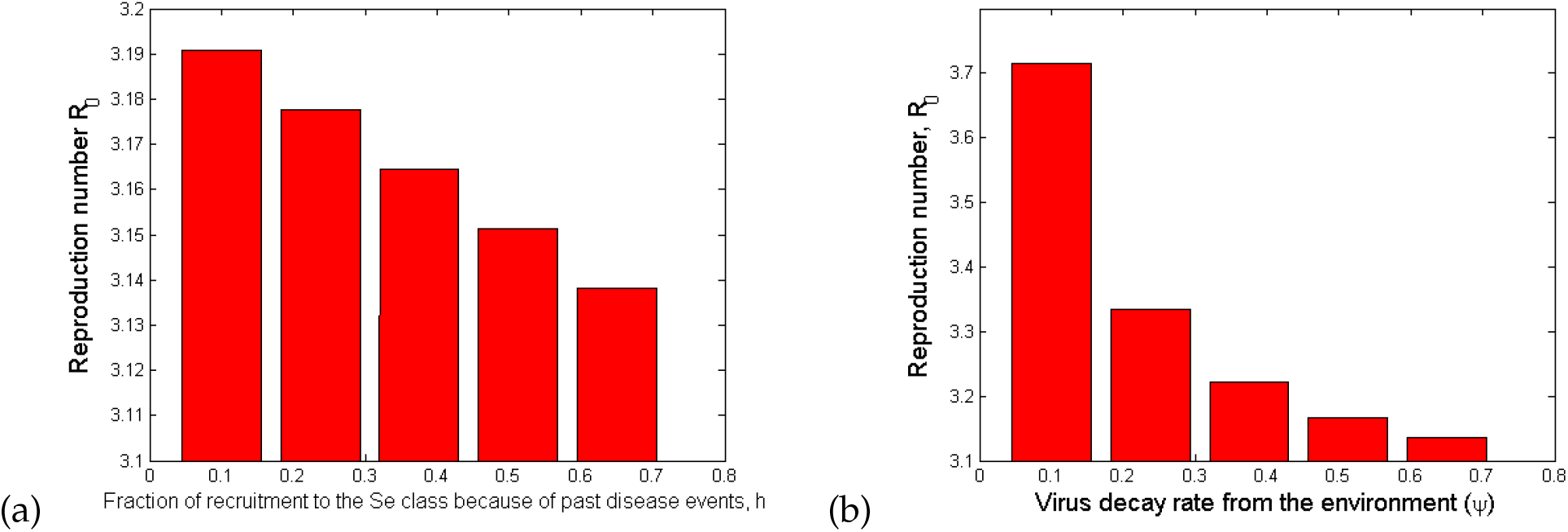
Bar plots showing the effect of the most sensitive parameters to ℛ_0_: (4(a)) Fraction of recruitment to the Se class because of past disease events; (4(b)) Virus decay rate from the environment. The values of the parameter values used are given in Table 3.

We observe in Figure 5 that the increase in person to person contact, *β*_1_ (Figure 5(a)), in poor personal hygiene, *β*_2_ (Figure 5(b)), and in the rate of shedding of the virus into the environment by both carriers (Figure 5(c)) and symptomatic individuals (Figure 5(d)) increases the value of ℛ_0_ and therefore the disease burden. It is evident that the most effective way of containing the infection is by minimizing contact, which is why most cases imposing a lockdown becomes an effective way of slowing the spread of the infection. In addition, good hygiene practices by all individuals are two-fold: (1) avoiding touching surfaces, always washing hands with soap and water, or using alcohol based hand sanitizer, which reduced the likelihood of contracting the pathogen from the environment; (2) those who are sick with symptoms like cough and flu, ought to use masks, when they cough or sneeze, must do so in a sanitary tissue which is then properly disposed off. We also note that hygienic practices without social/physical distancing may not significantly slow down the infection.

**Figure 5:**
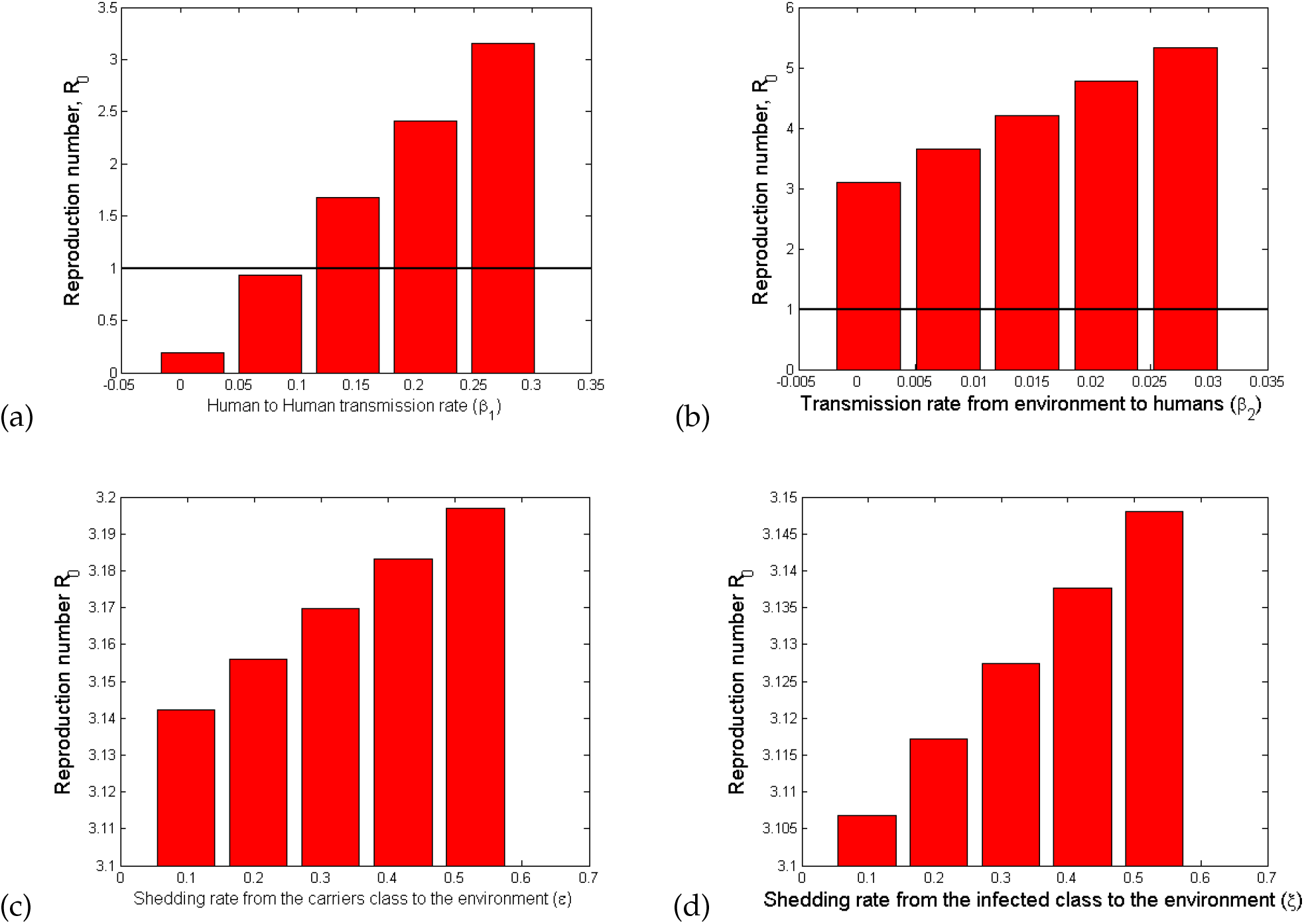
Bar plot showing the effect of the most sensitive parameters to ℛ_0_: (5(a)) Rate of disease transmission directly from human to human; (5(b)) Rate of disease transmission from the environment; (5(c)) Shedding rate from the carriers class to the environment (*ε*); (5(d)) Shedding rate from the infected class to the environment (*ξ*). The values of the parameter values used are given in Table 3.

In summary, we observe that it is possible to reduce the value of ℛ_0_ to a value less than unity by reducing only the value of *β*_1_ below 0.1 (see Figure 5(a)). This observation is in direct agreement with mitigation approaches that are aimed at minimising human-to-human contact (such as social distancing and imposing a lockdown). Therefore, the paremeter *β*_1_ is more influential in the model and can also play a significant role in eradication of the disease. The other parameters (see Figures 4, 5(b), 5(c) and 5(d)) may reduce the value of ℛ_0_ significantly when applied in combination but not as independent mitigation processes.

### 4.3 Numerical simulations and mitigation strategies

There are various ways of intervention mechanisms for COVID-19 that are observed being implemented in different part of the world. The strategies differ from country to country depending on the scientific information available to decision makers. For the simulation purpose of this study, we considered five different cases or scenarios of how to apply the interventions. The strategies described in each of the cases below are in addition to the awareness creation for voluntary self-protective mechanisms which are widely communicated through various media outlets. Here, we assume that the average effectiveness of the self-protective measures is 15% (as estimated from the data and reported in Table 3), and the individuals who decided to use any one of them are strict in following the appropriate rules.

**Case 1:** In this case, we assume that about 40% of the symptomatic infectious individuals and only 1% of the asymptomatic infectious individuals are detected and quarantined. This scenario is based on the assumption that among the people in the *I* class only about 40% show “above mild” symptoms and hence visit health care facilities, while the remaining individuals in this class (nearly 60% of them) remain at home or at large in the society. Then, through contact tracing mechanisms corresponding the hospitalized individuals, some people will be traced and tested, thereby about 1% of the total asymptomatic individuals can be detected.

A similar scenario is being applied currently in some sub-Saharan African countries.

The time profile in Figure 6 shows the situation described in Case 1. From this graph we can observe that the infection stabilizes around its endemic equilibrium, which is nearly at 5000 cases. (This number depends on the initial conditions and the demographic variables of the population under study.) This shows that the disease persists in the population.

**Figure 6:**
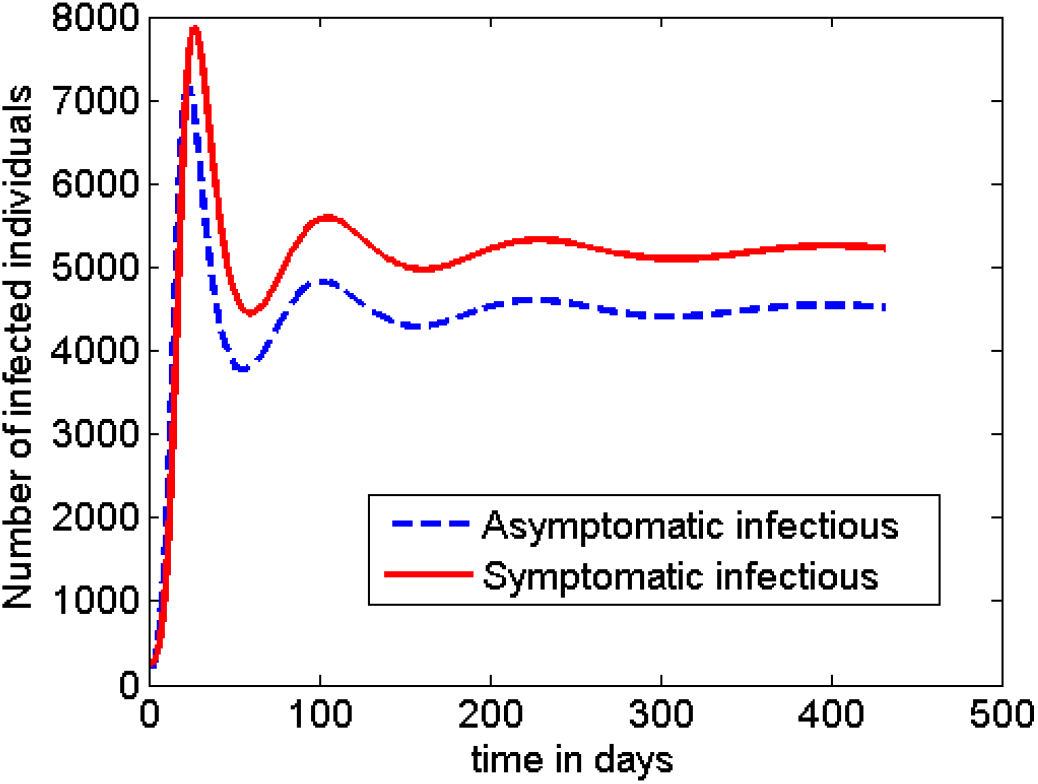
Dynamics of the disease with no additional intervention is applied.

**Case 2:** In this case, we assume that strict and longer time (6 weeks) of social distancing rules are enforced by the government nearly 4 weeks after the first positive case of COVID-19 is reported in the community.

We assume for simulation purpose that the implementation of the intervention strategy is divided into the following 4 time phases.

Phase 1: The first phase, in this case, is 24 days long (measured starting from the first positive case of COVID-19 is reported). During this phase because of lack of information and the nature of the infection, assume (as in Case 1) that only 40% of symptomatic infectious individuals and 1% of the asymptomatic infectious individuals are detected and quarantined.

phase 2: The second phase is assumed to last for 6 weeks (42 days) in this case. During this period, it is also assumed that

* 80% of the symptomatic class and 30% of the asymptomatic class are detected and quarantined,
* a mandatory social distancing rule is imposed, which is assumed to have a 70% reduction of effective contacts of individuals in the society,
* environmental disinfection is widely carried out, which is assumed to result in a 50% reduction in the rate of infection from the environment, and to contribute about the same percent impact in increasing the rate of decay of the pathogen from the environment.

Phase 3: The third phase is assumed to be 4 weeks (28 days) long, and is characterised by the partial lift of the ‘lockdown’ imposed in Phase 2. During this period, it is assumed further that

* 70% of the symptomatic class and 25% of the asymptomatic class are detected and quarantined,
* a relaxed social distancing rule is exercised, which is assumed to have a 25% reduction of effective contacts of individuals in the society,
* environmental disinfection is partially carried out, which is assumed to have an impact of reducing the rate of infection from the environment by 30% and increasing the rate of decay of the pathogen from the environment by the same 30%.

Phase 4: The last and fourth phase is the time when the social distancing rule is fully lifted. Due to the lesson learnt from the previous phases, we assume that the following interventions will continue during this period as well

* 70% of the *I* class and 10% of the *C* class are detected and quarantined,
* environmental disinfection is partially carried out, which is assumed to have an impact of reducing the rate of infection from the environment by 20% and increasing the rate of decay of the pathogen from the environment by the same 20%.

The time profile of the disease dynamics after implementing the above described interventions strategy is plotted in Figure 7. The figure shows that the count of the infected individuals decreases down to nearly zero in Phases 2 and 3, but the disease returns back into the society soon after. However, the peak of the second wave looks to be much more smaller than the first one. That means, the intervention mechanisms described in the above 4 phases of this case are not enough to contain the disease, and unless some additional intervention mechanisms are developed the disease persists in the society.

**Figure 7:**
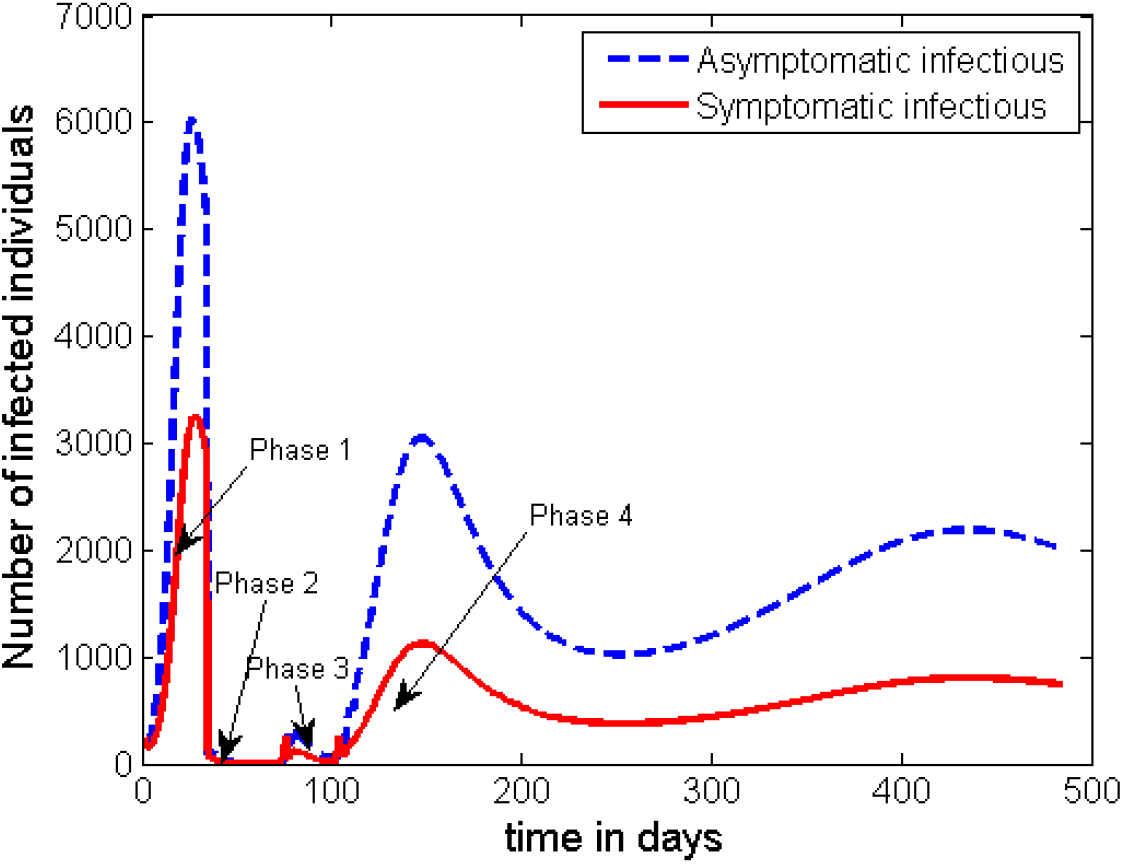
Dynamics of the disease with a mandatory 6-weeks lockdown and a 4-weeks of partial social distancing is imposed as described in Case 1.

**Case 3**: In this case, we assume that early action with shorter time social distancing rule is applied. In this scenario, it is assumed that the interventions described in Case 1 started half way through the time Phase 2 was implemented in Case 1. That means, the implementation of the interventions described in the four phases of Case 1 is assumed to be followed, but the length of the time in Phases 1 and 2 is reduced as described below.

1. Phase 1 lasts only 12 days,
2. Phase 2 lasts only 3 weeks, and
3. Phase 3 lasts 4 weeks (the same as in Case 2).

Otherwise all the details of the interventions in Case 1 are kept the same. The time profile for this set of interventions is given in Figure 8. The general behaviour of the graph in Figure 8 is the same as that of Figure 7. However, this strategy has an advantage in significantly reducing the height of the first peak. The height of the subsequent peaks are found to be the same unless some additional measures are taken after Phase 3.

**Figure 8:**
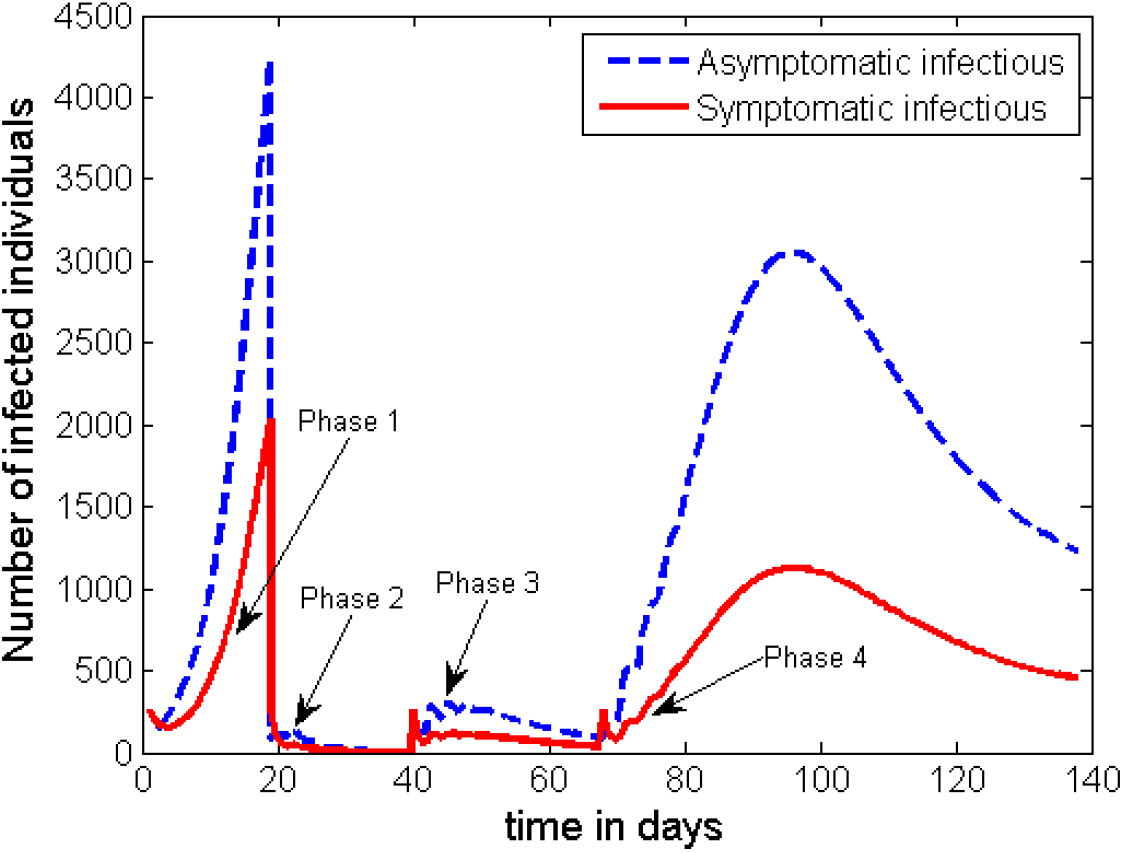
Dynamics of the disease (portraying the scenario in Case 2) with the start of early intervention measures but for half of the time as compared to that of Case 1.

Unfortunately the strategies in both of the above two scenarios (Case 2 and Case 3) do not help to fully contain the disease once it spreads in the population. As can be seen from Figures 7 and 8 another wave of outbreak of the disease will emerge at a later stage. Here, we can see that the asymptomatic infectious individuals play the greater role in becoming the major source for the second wave. Therefore, if there is a possibility to track and detect people with asymptomatic infection, and if they can be effectively quarantined for the required period of time there is a possibility for disease to be contained. As it can be observed from Figure 9, if we can increase the effect of detecting and quarantining the asymptomatic individuals to a proportion of about 30%, it is possible to significantly reduce the infection to a level that it cannot be a public treat. Otherwise, any lower proportion of this effort will imply the emergence of a second wave of infection in the community.

**Figure 9:**
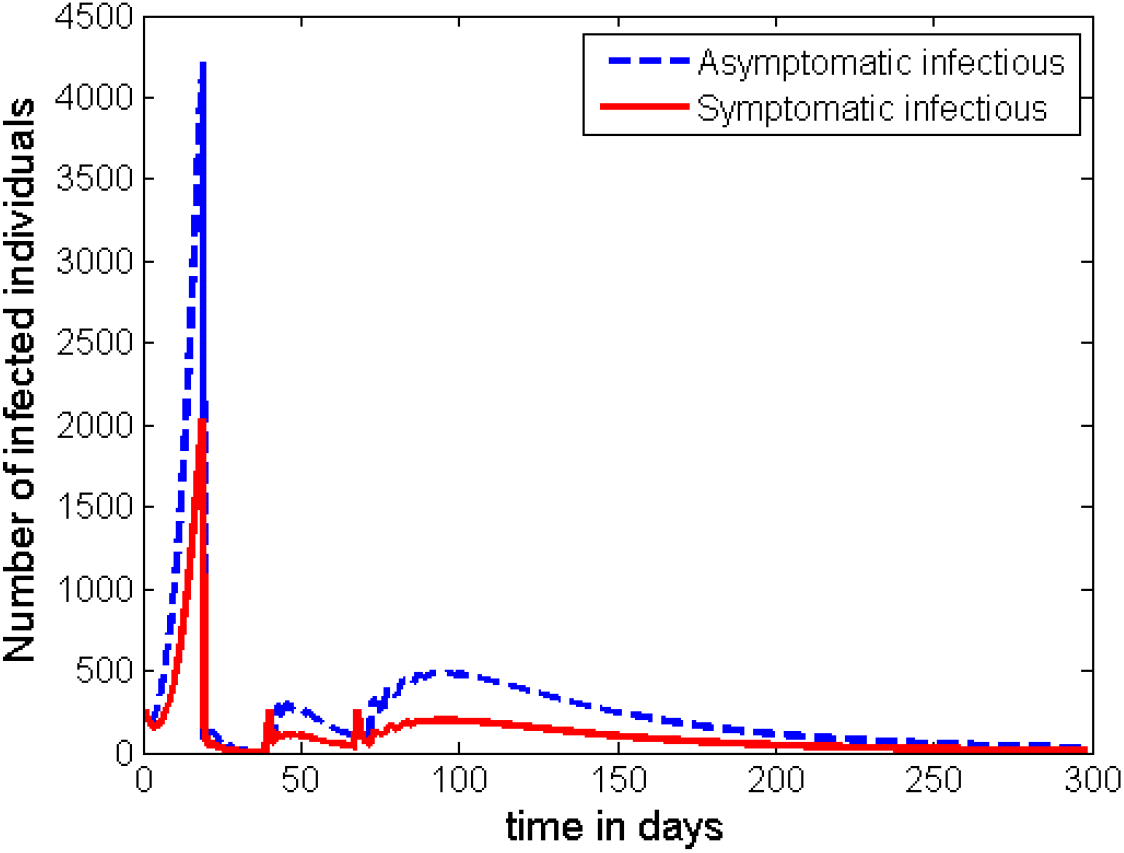
Dynamics of the disease with 30% of the *C* class and at least 70% of the *I* class are detected and quarantined in Phase 4 (after the interventions described in Case 2 are carried out).

Therefore, to contain COVID-19 in every given community, public health authorities need to work more on the detection and quarantining of the asymptomatic infectious individuals.

**Case 4**: In this case, we assume that no lockdown but only large number of testing is applied to detect and quarantine large proportion of infected cases.

If it is possible to beef up the effort of tracing the asymptomatic infectious individuals and be able to quarantine at least 35% of them continuously and effectively, our simulation shows that there is a possibility for the disease to be contained without imposing the strict lockdown rule on the total population. The plot in Figure 10 shows the time profile of the count of the infected groups while about 50% of the individuals from *I* class are effectively quarantined (for example inside appropriate health facilities).

**Figure 10:**
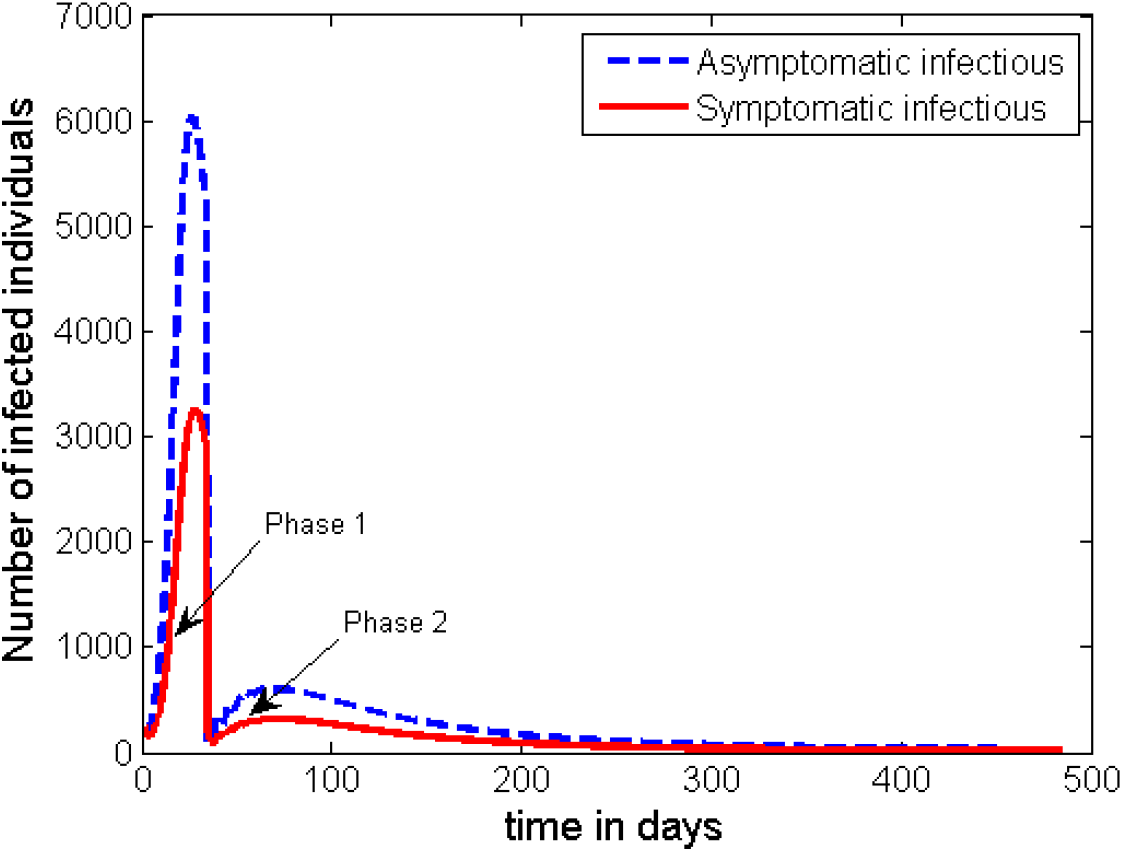
Dynamics of the disease with 30% of the population in the *C* class and at least 50% of the *I* class are detected and quarantined just after Phase 1 period.

We can observe that this intervention strategy can also produce the required result in containing the outbreak as some countries (like South Korea) is currently being following this pattern.

**Case 5**: In this case, we assume that the length of the lockdown period is nearly twice to the scenario in Case 2. But the effort in detecting the asymptomatic infectious individuals is kept minimum. This scenario is more applicable in highly resource constrained countries as the current cost of testing is high. In this case, it is assumed that the length of duration for each phase (except for Phase 2) is the same as given in Case 2. However, it is supposed that

1. the conditions in Phase 1 remains the same,
2. Phase 2 lasts 11 weeks with 50% of the symptomatic individuals and 5% of the asymptomatic individuals are detected and quarantined, while strict social distancing rule with an effect of reducing 70% of human contacts and 50% of environmental variables,
3. Phase 3 lasts 4 weeks (the same as in Case 2), but with 50% of individuals in the *I* class and 10% of individuals in the *C* class are detected and quarantined, while partial social distancing rule with an effect of reducing 25% of human contacts and 25% of environmental variables,
4. Phase 4 continues with detecting and quarantining 50% of members in the *I* class and 10% of members in the *C* class, while the other mandatory intervention are lifted.

The time profile of the infection following the scenario in Case 5 is plotted in Figure 11.

**Figure 11:**
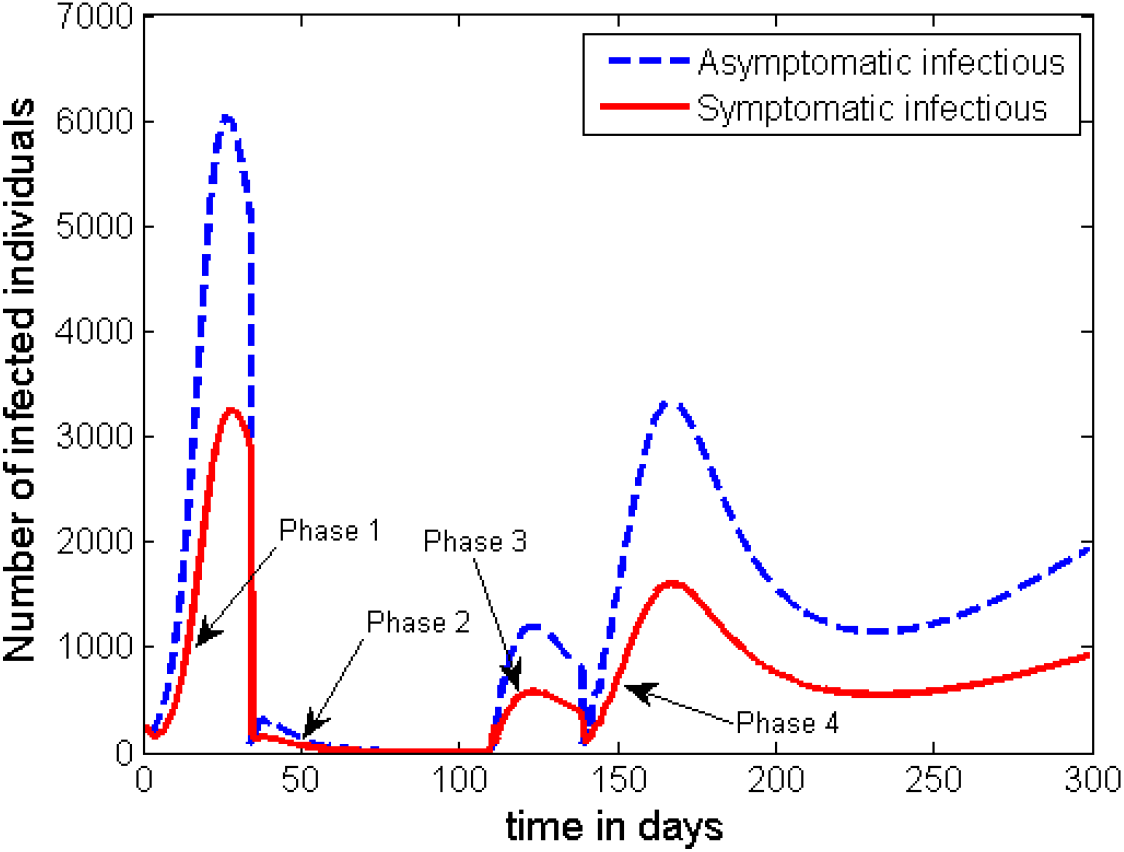
Dynamics of the disease with at most 10% of the population in the *C* class and at least 50% of the *I* class are detected and quarantined just after Phase 1 period, with strict social distancing rule imposed for 11 weeks.

The simulation out for this scenario shows that even if we increase the length of lockdown period to 11 weeks (like it was practised in the Hubei province, China) the disease may re-emerge after some period of time. However, the heights of the peaks in the subsequent waves of the disease are much more less than that of the first peak. Therefore, once again, unless the authorities apply some kind of strict contact tracing mechanism and conduct enough testing to detect and isolate up to 30% of the asymptomatic infectious individuals, the disease persist in the community with multiple subsequent waves.

In general, from the simulations, we can observe that in all of the above scenarios a transition from one phase to the other intervention phase is characterised by a surge in new cases. However, the number will eventually go down if the intervention in the immediate next phase is effective, and otherwise the disease re-emerges in the population.

## 5 Conclusions

We presented a mathematical model for the dynamics of COVID-19 whose first cases were reported in December 2019 in Wuhan-China. The model incorporates a behaviour change function to account for proportion of individuals who decided to use any of the self-protective measures and adhere to it. In addition, it also considers a proportion of individuals with history/knowledge of similar infections from the past and practice necessary protective measures right from the beginning. The model also accounts for asymptomatic carriers of the infection as well as the concentration of the pathogen within the environment. The basic properties of the model including well-posedness, the disease free equilibrium and its stability, model basic reproduction number as well as existence of backward bifurcation were examined. To estimate the parameter values, the model was fitted to the data on daily new cases reported in WHO situation reports 1-57 [43], which accounts for the period from January 21, 2020 to March 17, 2020. From the nominal values from the data fitting, we obtained a reproduction number, ℛ_0_ ≈ 2.9 (2.1-4) which compares well with the values of ℛ_0_ obtained in other researched, for instance, (2.24 − 3.58) [51] and 3.11(95%CI, 2.39 − 4.13) [34]. From our sensitivity analysis simulations, we observed that for some given parameter combinations the values of ℛ_0_ can be reduced to below 1, and similarly to values much higher than 4.

We observed that if the recovering individuals do so with permanent immunity (*ϕ* = 0), then reducing the reproduction number to a value below unity is enough to contain the infection. On the other hand, if recovering individuals do so with temporary immunity (*ϕ* ≠ 0), the proposed model exhibits backward bifurcation, which implies that reducing the value of ℛ_0_ below 1 is not enough to contain the infection.

By applying the Latin Hypercube sampling scheme, we observed that if the disease is to be easily contained, measures such as; physical/social distancing (which reduces the rate of disease transmission directly from human to human), improved personal hygiene (which reduces the rate of disease transmission from the environment to humans), and minimal shedding of the pathogen into the environment by both asymptomatic and symptomatic individuals, have the greatest potential of slowing the epidemic. We further observed that increased decay of the pathogen from the environment (achieved by disinfecting surfaces) is less significant in reducing/curbing the number of cases.

We further observed that having high numbers of people with knowledge from previous similar infections, that are practising the prescribed self-protective measures can delay/slow down the otherwise potentially explosive outbreak. Consequently, the daily number of cases is kept at low manageable levels. In addition, increasing the average effectiveness of the self-protective measures and adherence to such measures is vital in realising low peaks of the number of cases. Furthermore, due to the absence of vaccination or any approved medication, developing capacity to detect carrier groups is very important. From our results, it is recommended that countries should develop capacities to identify and quarantine at least 30% of carriers as well as at least 50% of symptomatic cases if the infection is to be controlled. Our model predicts a possible resurgence of the number of cases if the asymptomatic cases are still many by the time disease spread curbs/lockdown measures are lifted. It is also evident that practising social/physical distancing, good hygiene and disinfecting surfaces to eliminate the virus from surface are vital in reducing the disease burden. But the contribution of disinfecting surfaces is not that high. In addition, we observe from simulations that although disease spread curbs (such as a lockdown measure) may be imposed, their real impact on the number of new cases may only be realised after approximately 21 days, and the reduction (when it appears) could be sharp in the case of a strict lockdown measure with high impact in reducing effective contact between individuals in the population.

When providing the mitigation strategies, we did not account for the delay between the actual incidence and the point when cases are confirmed since the actual parameters describing such a delay are not known. In health systems where testing of suspected cases is done after individuals show symptoms or on demand, it is likely to have a big gap between the actual incidence and confirmation of new cases. The impact of the delay between actual incidence and confirmation of case can be explored in future work. In addition, we assumed that all individuals who recover, do so with the same level of immunity. However, this may not necessarily be the case since immunity of individuals is affected by a number of factors including age, cortisol levels and nutrition among others. The impact of differentiated levels of immunity on the disease dynamics and potential resurgence of the epidemic can be explored in the future when relevant data becomes available. Our model did not include the possibility of vaccination or treatment. We however acknowledge their importance in controlling the infection. Therefore, optimal control of infection in presence of these mitigation strategies can be explored in the as some of the relevant data become available.

## Data Availability

Data is available from the WHO situation reports 1-57
https://www.who.int/emergencies/diseases/novel-coronavirus-2019/situation-reports

## Acknowledgments

The authors S.M.K. and H.J.B.N. gratefully acknowledge Botswana International University of Science and Technology (BIUST) that supported their research through the project entitled *‘Research Initiation grant of the office of the DVCRI of BIUST, with grant number DVC/RDI/2/1/161(34)*.

## Appendix A Proof of Theorem 3.1

The proof of Theorem 3.1 is outlined here below based on the following two steps.

First, we show that all solutions of Eq. (2.2) are nonnegative as required in [3, 37]. To show that the state variables *S* and *S*_*e*_ of the model are positive for all *t* ≥ 0, we use proof by contradiction. We suppose that a trajectory crosses one of the positive cones at times *t*_1_ or *t*_2_ such that:

- *t*_1_: *S*(*t*_1_) = 0, *S*t(*t*_1_) < 0, *S*_*e*_(*t*) > 0, *C*(*t*) > 0, *I*(*t*) > 0, *R*(*t*) > 0, and *E*(*t*) > 0 for *t* ∈ (0, *t*_1_), or
- *t*_2_: *S*_*e*_(*t*_2_) = 0, 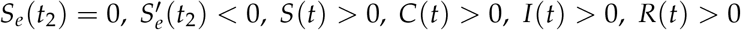, and *E*(*t*) > 0 for *t* ∈ (0, *t*_2_),

Using the first equation of Eq. (2.2), the first assumption leads to

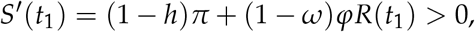

which contradicts the first assumption that *S*′(*t*_1_) < 0. Thus, *S*(*t*) remains positive for all *t* ≥ 0. Here, *t*_1_ is chosen so that our point to be on the positive axis of *S*(*t*) so that *R*(*t*_1_) is positive.

Using the second equation of Eq. (2.2),

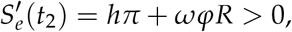

which also contradicts the assumption 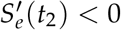. Hence, *S*_*e*_(*t*) remains positive for all *t* ≥ 0. Based on the third equation of Eq. (2.2),

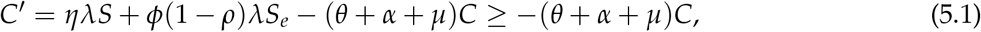

because *S*(*t*) and *S*_*e*_(*t*) are nonnegative for *t* ≥ 0. Solving Eq. (5.1) yields

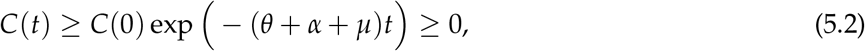

Likewise, from the fourth equation of (2.2), we obtain

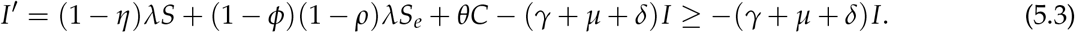

Solving (5.3) leads to

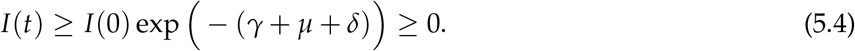

Similarly, using the last two equations of Eq. (2.2), we have

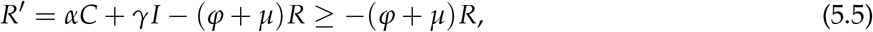

and

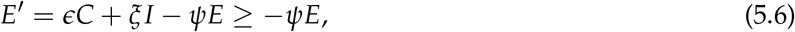

because *S*(*t*), *S*_*e*_(*t*), *C*(*t*), and *I*(*t*) are nonnegative for *t* ≥ 0. Solving Eqs. (5.5) and (5.6) gives

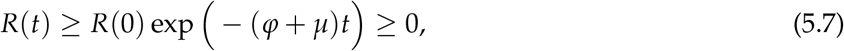

and

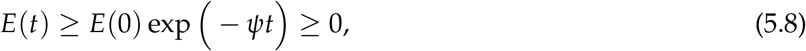

respectively.

Thus, any solution of Eq. (2.2) is nonnegative for *t* ≥ 0 and any initial condition in Ω. Finally, the total number of the population *N*(*t*) at time *t* is governed by

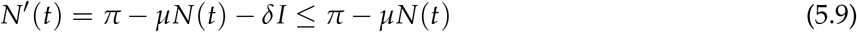

Thus, for the initial data 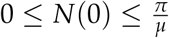, by Gronwall inequality, we obtain

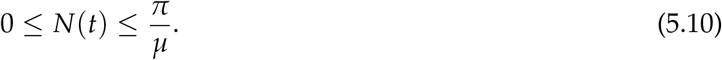

Moreover, for the environmental variable *E*′ we have

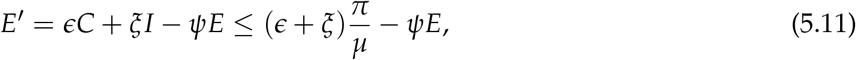

because *C*(*t*) and *I*(*t*) are less than 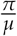 for all *t* ≥ 0. Applying again the Gronwall inequality, for 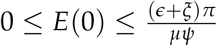, leads into

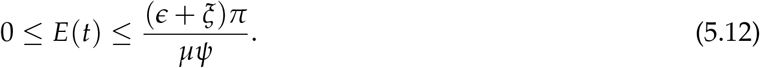

Combining the above two steps and Theorem 2.1.5 in [11] for the existence of unique bounded solution, we infer that any solution of Eq. (2.2) is nonnegative and bounded. Hence, Eq. (2.2) defines a dynamical system on Ω. □

## Appendix B: Proof of Theorem 3.3

**Proof:** The theorem is the direct application of Theorem 4.1 in [6]. To check the existence of backward bifurcation of the model Eq. (2.2) at ℛ_0_ = 1, we use the center manifold theorem [6]. For this purpose, we introduce the following change of variables.

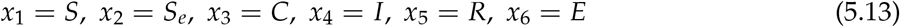

so that

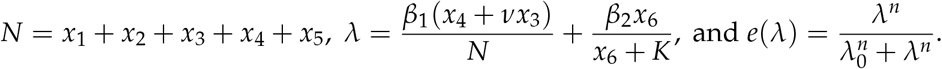

Moreover, by using the vector notation *X* = (*x*_1_, *x*_2_, *x*_3_, *x*_4_, *x*_5_, *x*_6_)^*T*^, the model Eq. (2.2) can be written in the form *X*′ (*t*) = *F* = (*f*_1_, *f*_2_, *f*_3_, *f*_4_, *f*_5_, *f*_6_)^*T*^ as follows:

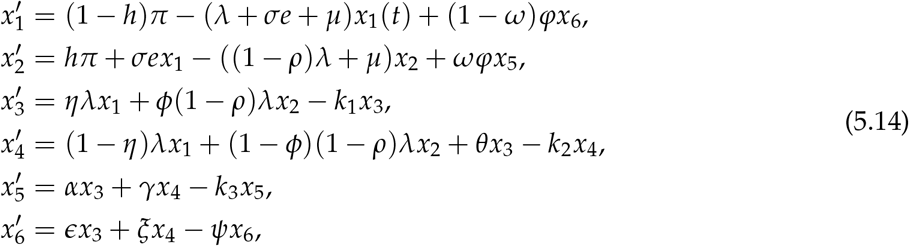

where,

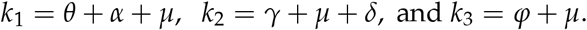

When ℛ_0_ = 1 and *β*_1_ is considered as a bifurcation parameter, from (3.5) we get

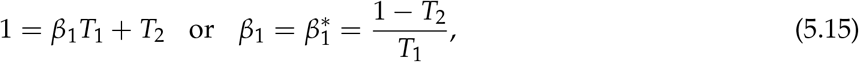

Where

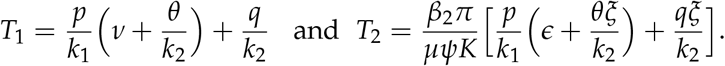

Further more 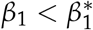 if and only if ℛ_0_ < 1 and 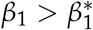 whenever ℛ_0_ > 1.

The Jacobian of the system (5.14) at the associated *DFE* (*ε*_0_) is

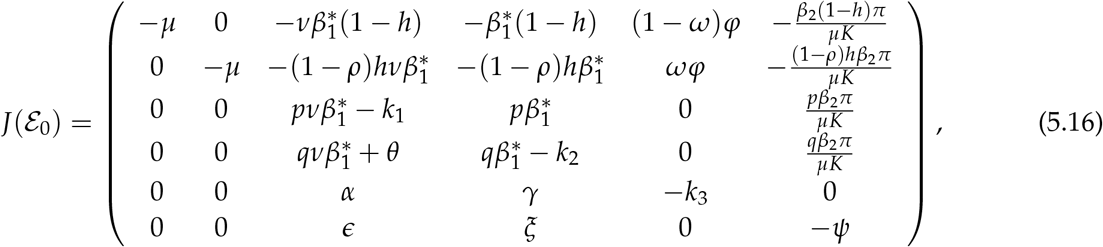

where *p* = *η*(1 − *h*) + *ϕ*(1 − *ρ*)*h*, and *q* = (1 − *η*)(1 − *h*) + (1 − *ϕ*)(1 − *ρ*)*h*.

The transformed system Eq. (5.14), with 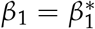, has a non-hyperbolic equilibrium point such that the linear system has a simple eigenvalue with zero real part and all other eigenvalues have negative real parts. Hence, the centre manifold theory [6] can be used to analyse the dynamics of the model Eq. (5.14) near 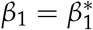. By using the notation in [6], the following computations are carried out.

The right-eigenvector

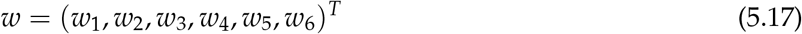

associated with the zero eigenvalue of *J*(E_0_) such that

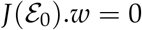

at 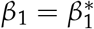 is given by

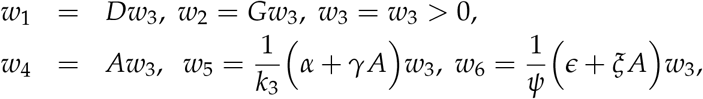

where

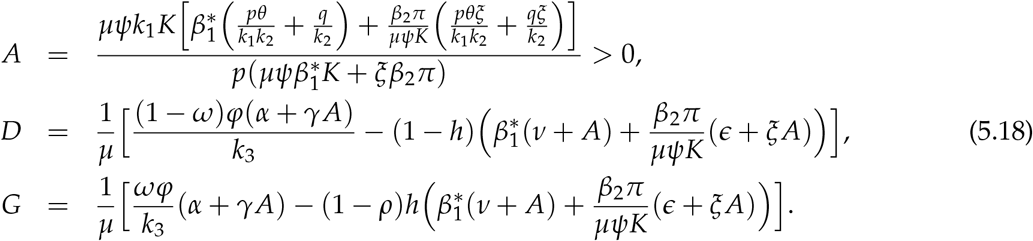

Similarly, the left-eigenvector

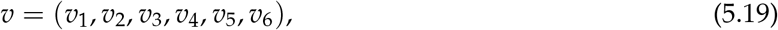

of *J*(*x*∗) such that

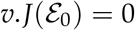

associated with the zero eigenvalue is given by,

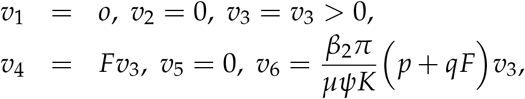

Where

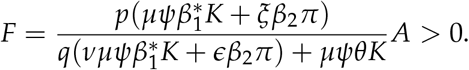

The right-eigenvector *w* and the left-eigenvector *v* need to satisfy the condition *v*.*w* = 1.

The bifurcation coefficient *a* at the DFE (*ε*_0_) is given by

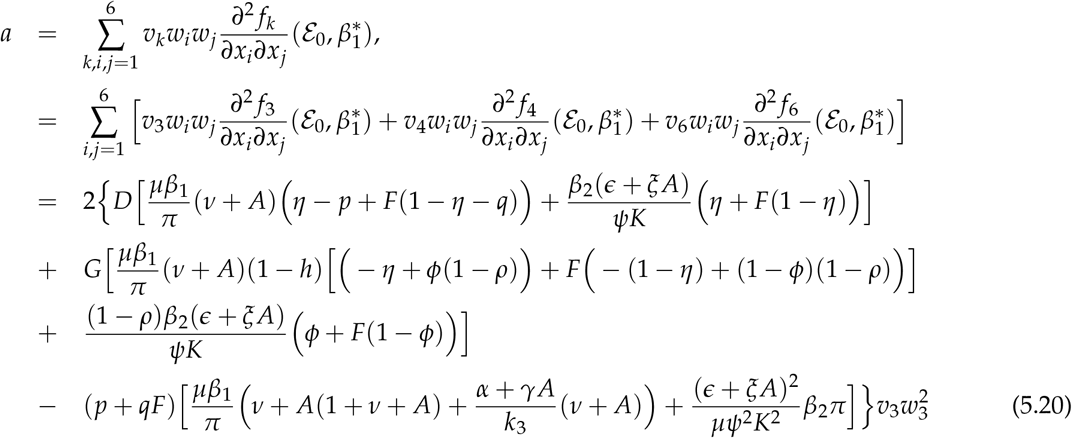

Thus, the bifurcation coefficient *a*, can be positive for the right choice of the parametric values that satisfy the condition in Eq. (3.6).

The second bifurcation coefficient *b* is given by

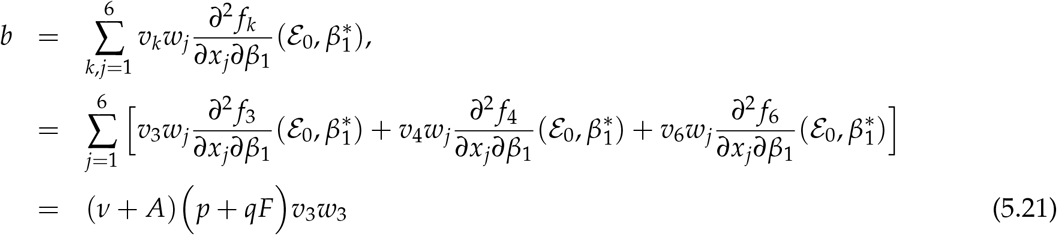

Clearly, *b* > 0 because *A* and *F* are positive.

When *ϕ* = 0, *D* and *G* in (5.18) are negative and *a* in (5.20) is negative as well. Hence, by Theorem 4.1 in [6], the model will not exhibit a backward bifurcation at ℛ_0_ = 1.

